# Cost-Effectiveness of Sulbactam-durlobactam Compared with Colistin for Treating Carbapenem-Resistant *Acinetobacter baumannii* Infections in Critical Care Settings in the United States

**DOI:** 10.64898/2026.09.13.26362951

**Authors:** A J Ologunowa, A R Caffrey, S A Cohen, S Kogut

## Abstract

**Objective:** Carbapenem-resistant *Acinetobacter baumannii* (CRAB) infections present a significant public health challenge in the United States due to multidrug resistance, substantial added inpatient costs, and subsequently high mortality rates.

**Method:** A decision-analytic model was developed to evaluate the cost-effectiveness of sulbactam-durlobactam compared to colistin for treating CRAB infections, using data from the Acinetobacter Treatment Trial Against Colistin (ATTACK) trial, in which patients received either sulbactam-durlobactam or colistin alongside imipenem-cilastatin. The model incorporated survival rates, clinical outcomes (complete or incomplete cure), and the incidence and costs of acute kidney injury, which is more frequent in colistin-treated patients. Deterministic sensitivity analysis and two scenario analyses were performed.

**Results:** The base case analysis estimated an expected per-patient cost of $57,887 for sulbactam-durlobactam compared to $55,157 for colistin. Patients treated with sulbactam-durlobactam had higher survival than those treated with colistin (0.66 versus 0.46), corresponding to 0.19 additional life saved per patient. The incremental cost-effectiveness ratio (ICER) for sulbactam-durlobactam compared with colistin was $14,097 per additional life saved. The ICER was most sensitive to the proportion of survivors with incomplete clinical cure for both treatments, the costs of sulbactam-durlobactam and cefiderocol, and survival outcomes in patients treated with sulbactam-durlobactam. Scenario analyses showed that the cost-effectiveness of sulbactam-durlobactam was sensitive to clinical cure outcomes, with ICERs ranging from $69,758 per additional life saved to dominance favoring sulbactam-durlobactam under different assumptions.

**Conclusions:** Although treatment costs were higher, sulbactam-durlobactam has a favorable cost-effectiveness profile as compared with colistin when accounting for the costs of managing acute kidney injury.

## Introduction

Carbapenem-resistant *Acinetobacter baumannii* (CRAB) is classified by the Centers for Disease Control and Prevention (CDC) as an urgent public health threat in the United States (US) due to limited treatment options and significant healthcare burdens.^1^ CRAB infections primarily affect vulnerable patients with a high comorbidity burden and those who are immunocompromised.^2^ These infections increase the duration and cost of hospitalizations, particularly in critical care, where patients are more susceptible to severe complications and increased mortality.^2,3^ Multidrug-resistant *Acinetobacter baumannii* causes 80% of hospital-acquired and ventilator-associated pneumonia globally, and 70% of such infections in the US.^4,5^ Despite modest reductions in laboratory-identified CRAB isolates and CRAB-associated hospitalizations, CRAB remains a significant concern in critical care settings due to its high fatality rate.^6^ The attributable annual direct healthcare costs of CRAB infections in the US were estimated to be $281 million in 2017.^1^

According to the 2024 Infectious Disease Society of America (IDSA) treatment guidelines for CRAB infections, combination therapy with at least two agents is recommended, but there is no stated consensus on the standard of care antibiotic regimen.^7^ First-line antibiotic therapy options include ampicillin-sulbactam, tigecycline, colistin, and meropenem or imipenem-cilastatin, while cefiderocol is suggested to be limited to CRAB infections that have developed resistance to other first-line antibiotics.^7^

Prior economic evaluations have examined the cost-effectiveness of newer β-lactam/β-lactamase inhibitor therapies for the treatment of CRAB infections compared with colistin.^8,9^ These studies have shown that when incorporating the costs and outcomes of drug-induced kidney injury the new beta-lactamase inhibitors (ceftolozane-tazobactam, ceftazidime-avibactam, meropenem-vaborbactam, and imipenem-relebactam) are cost-effective for treating CRAB infection compared to colistin, with incremental cost-effectiveness ratios of approximately $3,180 and $3,900 per quality-adjusted life-year (QALY).^8,9^ More recently, the Acinetobacter Treatment Trial Against Colistin (ATTACK) [NCT03894046],^10^ a phase 3, pathogen-specific randomized clinical trial, evaluated the efficacy of sulbactam-durlobactam compared with colistin, with both regimens administered in combination with imipenem-cilastatin and nephrotoxicity assessed as a safety outcome.^11^ The trial demonstrated improved clinical outcomes and reduced nephrotoxicity with sulbactam-durlobactam compared with colistin, particularly among critically ill patients. Using a decision-analytic model informed by data from the ATTACK trial, we evaluated the cost difference and cost-effectiveness of sulbactam-durlobactam compared with colistin in the inpatient critical care setting.

## Methods

### Model Overview

A decision-analytic model was developed to evaluate the cost-effectiveness of sulbactam-durlobactam compared with colistin for treating CRAB infections in critical care settings in the US, using efficacy data from the ATTACK study (NCT03894046),^10,11^ which found sulbactam-durlobactam to be non-inferior to colistin for all-cause mortality, which was higher in patients treated with colistin (32.3%) compared with sulbactam-durlobactam (19.0%).

The decision model incorporated survival, probability of survival with clinical cure or requiring the use of additional gram-negative antimicrobial therapy, and the incidence and costs of acute kidney injury (AKI), which was higher for patients treated with colistin. AKI outcome was stratified as risk, injury, failure, loss, and end-stage renal disease (RIFLE) criteria.^12^ Due to lack of information about the influence of kidney injury on mortality, survival was based solely on CRAB outcomes reported in the ATTACK trial. According to IDSA guidelines, cefiderocol is recommended to be reserved for cases where other antibiotics indicated for CRAB infections have proven ineffective.^7^ We assumed that patients in both treatment arms used similar background combination therapy with similar costs, and that survival differences were due to the treatment for CRAB infection. The base case values for the model parameters are presented in Table 1, and the model structure is illustrated in Figure 1.

**Figure 1:**
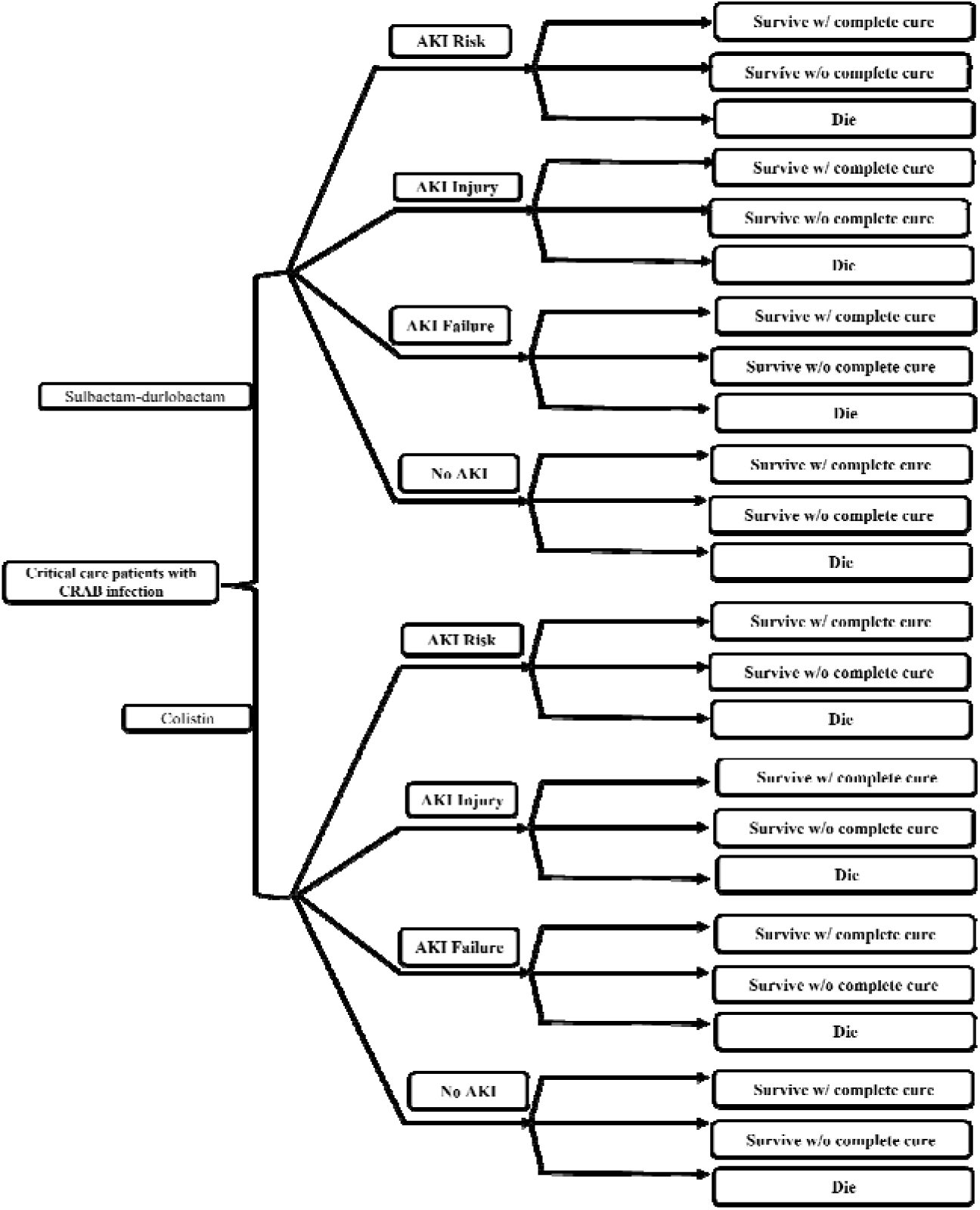
Decision tree for the base case. CRAB, carbapenem-resistant *Acinetobacter baumannii*; AKI, acute kidney injury

**Table 1:**
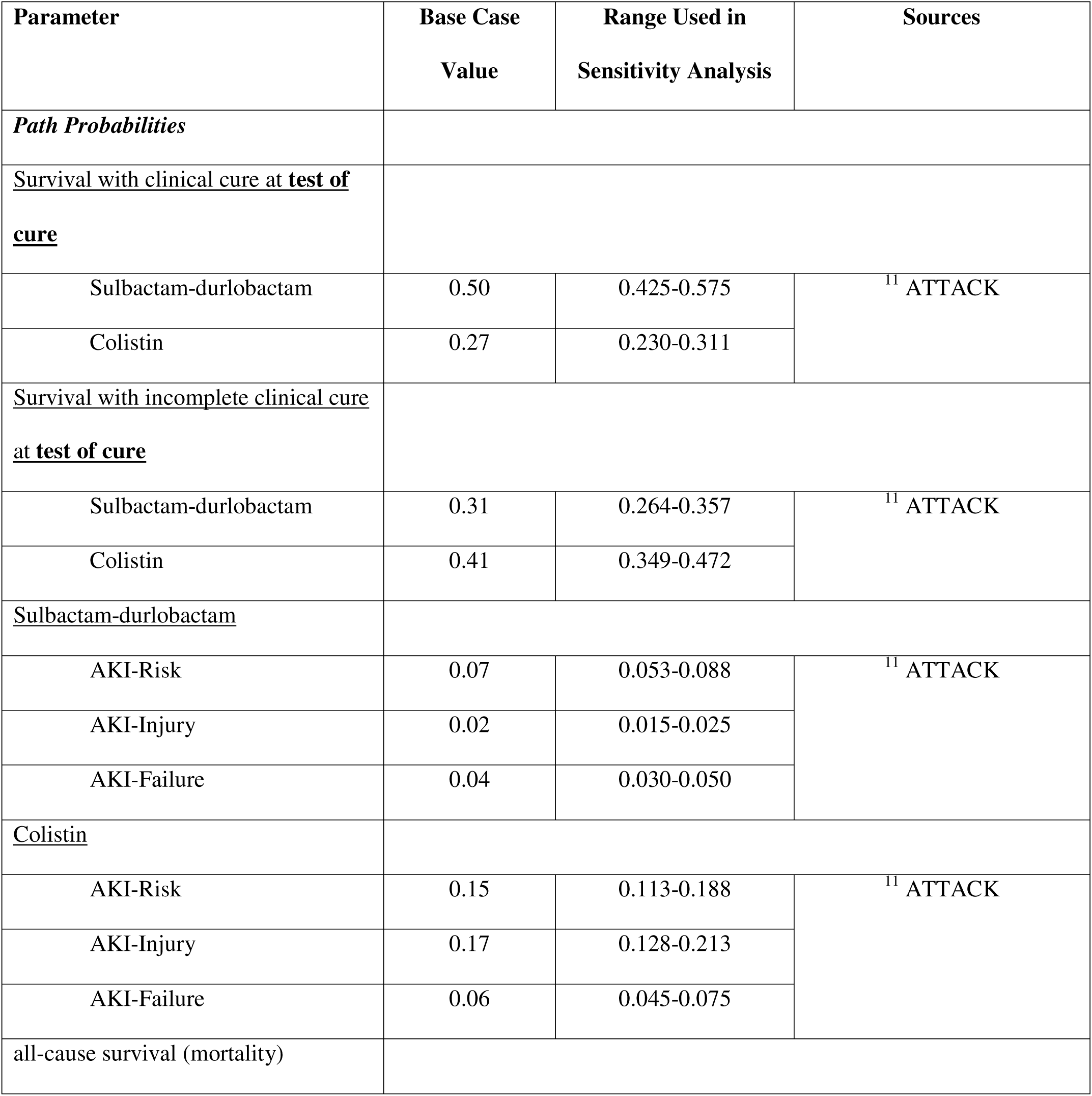
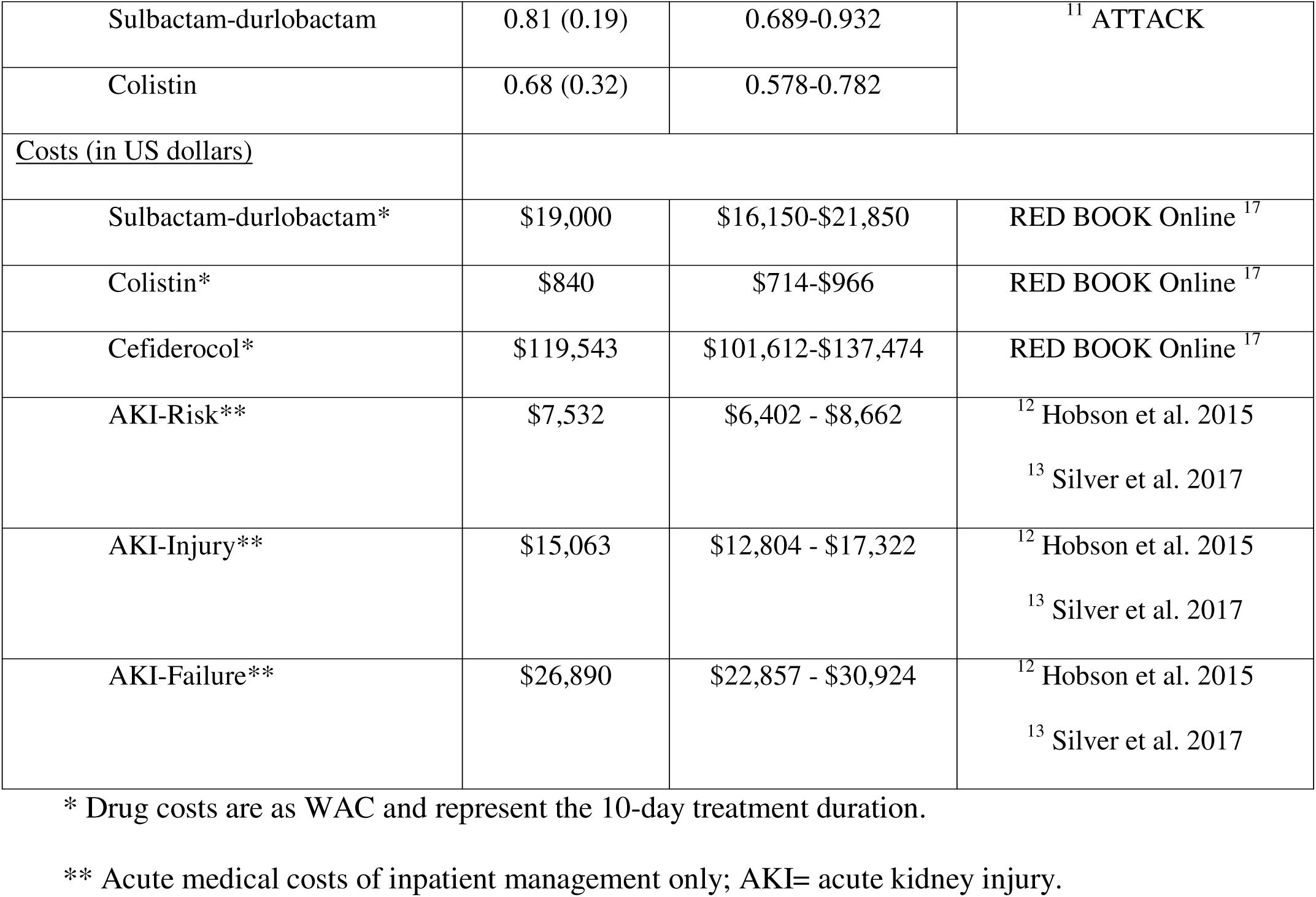
Clinical outcomes and medical costs.

### Costs

The model incorporated direct medical costs in the critical care setting, comprising Wholesale Acquisition Costs for sulbactam-durlobactam and colistin, and the cost of managing drug-related AKI. The 2024 IDSA CRAB treatment guidelines recommend a combination of sulbactam-durlobactam or colistin with at least one other antibiotic.^7^ Therefore, our study assumed that the cost and duration of use of background combination therapy for treating CRAB infections in critical care settings were similar in both groups, and as such, these offsetting costs were not included in the model. Recommended drug dosing was obtained from the Food and Drug Administration (FDA) label for sulbactam-durlobactam, and the daily dose was multiplied by 10 days to yield the treatment cost for the base case. Administration costs, intravenous supplies, laboratory monitoring, bed days, and other hospitalization costs were assumed to be similar for each treatment and were thus not included in the model. Silver *et al* (2017) and Hobson *et al*. (2015) reported the RIFLE-stratified cost of AKI as the risk-adjusted average incremental cost of care for patients with postoperative AKI compared to those without AKI over the follow-up period.^12,13^ These estimates were prorated for the relevant model branch durations. All costs were adjusted to 2025 US dollars using the consumer price index (CPI-U), except for medication, which was based on 2025 Wholesale Acquisition Cost (WAC).

### Outcomes

The model used a 45-day time horizon, consistent with the treatment and follow-up period of the ATTACK trial, in which patients with confirmed CRAB infections received either sulbactam-durlobactam or colistin, both in combination with imipenem-cilastatin, for 7 to 14 days, followed by 14 days of post-therapy follow-up. A mean treatment duration of 10 days was applied in the base case, with a range of 7 to 14 days explored in the deterministic sensitivity analysis; treatment duration was incorporated into the model to estimate per-patient drug costs, which were calculated on a per-day basis. Patients with incomplete clinical cure during the outcomes assessment period were assumed to transition to cefiderocol for a similar treatment duration as the next preferred agent in both the colistin and sulbactam-durlobactam arms in the model.

Based on the ATTACK trial, clinical cure was defined as complete resolution or significant improvement of confirmed CRAB infection without the need for additional gram-negative antimicrobial therapy, and incomplete clinical cure was defined as its complement (1 − clinical cure proportion). Survival with clinical cure and survival with incomplete clinical cure were derived by multiplying the respective cure proportions by the 28-day all-cause survival proportion. AKI outcomes were assessed within 30 days of the end of therapy. The proportion achieving clinical cure, incorporated as a path probability in the decision model, was assessed at the test-of-cure assessment 7 days after the end of therapy in the base case, clinical cure at the end of therapy in the first scenario analysis, and clinical cure at late follow-up 14 days after the end of therapy in the second scenario analysis.

Outcomes assessed included per-patient treatment costs and additional lives saved. The incremental cost-effectiveness ratio (ICER) was calculated as the expected per-patient incremental cost per additional life saved with sulbactam-durlobactam as compared to colistin. The ICER was compared to a willingness-to-pay (WTP) threshold of $81,624 (2025-adjusted value), derived from *Nelson et al.*, who estimated the attributable inpatient cost of invasive infections caused by carbapenem-resistant Acinetobacter species at $62,396 (2017 USD) using Veterans Affairs critical care data in the United States. This threshold, defined as the incremental inpatient cost attributable to infection, calculated as the difference in hospitalization costs between patients with positive cultures and matched controls adjusted for relevant confounders, was selected as the most contextually appropriate benchmark given the comparable US inpatient setting and patient population.^14^

### Sensitivity and Scenario Analysis

One-way deterministic sensitivity analysis (DSA) of input parameters was performed, using a Tornado diagram to visualize the influence of varying input ranges. The parameters included in the DSA were the probabilities of surviving with or without complete cure, incidence of each RIFLE-stratified AKI subcategory, and treatment costs. Each variable was systematically tested within the ranges specified in Table 1. For inputs without empirical ranges, we varied the input estimate by ± 25%, except for survival outcomes and costs, which were varied by +/-15% and +/-50%, respectively, in association with plausible ranges.^15^

We conducted two scenario analyses to examine the impact of parameter uncertainty and model structure on the base-case conclusions. The base case defined clinical response by cure status at the test-of-cure assessment conducted 7 days after therapy completion. In the first scenario analysis, clinical response was defined by complete or incomplete clinical cure at the end of therapy. In the second scenario analysis, clinical response was defined by complete or incomplete clinical cure at late follow-up. AKI risks were held at their base-case values in both scenarios. We used TreeAge Pro 2024 (TreeAge Software, LLC, Williamstown, MA, USA) and MS Excel for the decision models and analyses. Methods and results were reported according to the Consolidated Health Economic Evaluation Reporting Standards (CHEERS) reporting guideline.^16^

## Results

### Base Case Analysis

Base-case drug costs were $19,000 for sulbactam-durlobactam, $840 for colistin, and $119,543 for cefiderocol (US WAC, 10-day treatment duration). The incremental costs of managing AKI in the critical care setting were $7,532 for risk, $15,063 for injury, and $26,890 for failure. The estimated per-patient total expected cost per treatment episode, including drug acquisition and acute kidney injury management, was $57,887 for sulbactam–durlobactam and $55,157 for colistin. Patients in the model treated with sulbactam-durlobactam experienced greater survival than those treated with colistin (0.66 versus 0.46), corresponding to 0.19 additional life saved per patient. The ICER for sulbactam–durlobactam versus colistin was $14,097 per additional life saved (Table 2). The cost-effectiveness plot for the base case is shown in Figure 2.

**Figure 2:**
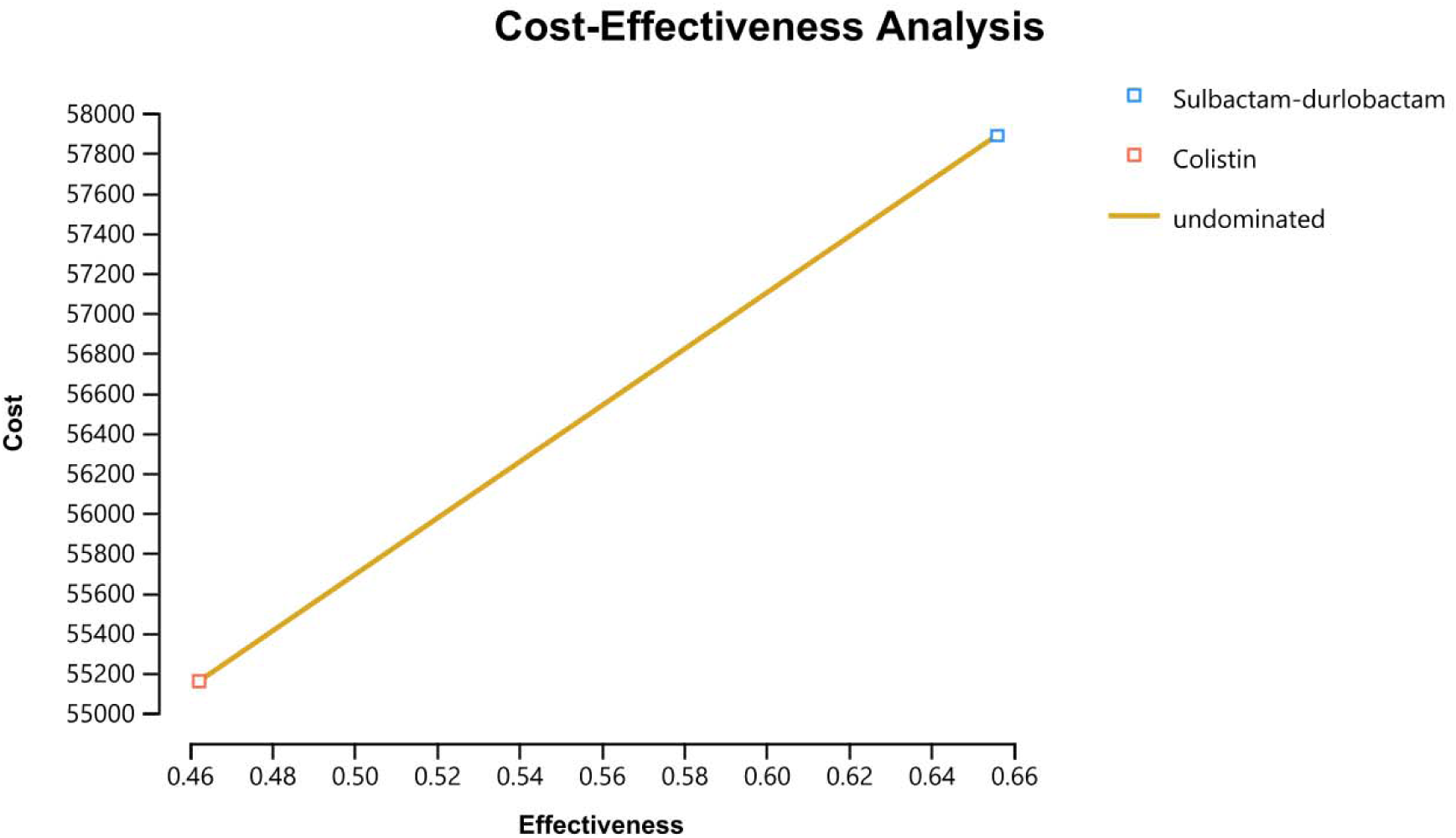
Cost-Effectiveness Analysis Plot for the Base Case (per additional life saved)

**Table 2:** Base case outcomes for sulbactam-durlobactam compared to colistin therapy.

| Strategy | Colistin | Sulbactam-durlobactam | Incremental<sup>+</sup> |
| --- | --- | --- | --- |
| 10-day drug cost | \$840 | \$19,000 | |
| Total costs | \$55,157 | \$57,887 | \$2,731 |
| Survival outcome | 0.46 | 0.66 | 0.194 |
| Incremental cost per additional life saved | | | \$14,097 |

<sup>+</sup>Incremental values were calculated from unrounded estimates and may not equal the difference of the rounded values shown.

### Sensitivity Analysis

The results of the deterministic sensitivity analysis are presented in the Tornado diagram (Figure 3). The ICER was influenced by variations in the proportion of colistin-treated patients surviving with incomplete clinical cure (-$30,890 to $42,617 per additional life saved), proportion of patients treated with sulbactam-durlobactam with incomplete clinical cure (-$17,696 to $36,023), cost of sulbactam-durlobactam (-$617 to $28,810 per additional life saved) and cefiderocol (-$4,840 to $23,354 per additional life saved), and survival among patients treated with sulbactam-durlobactam ($9,335 to $28,536).

**Figure 3:**
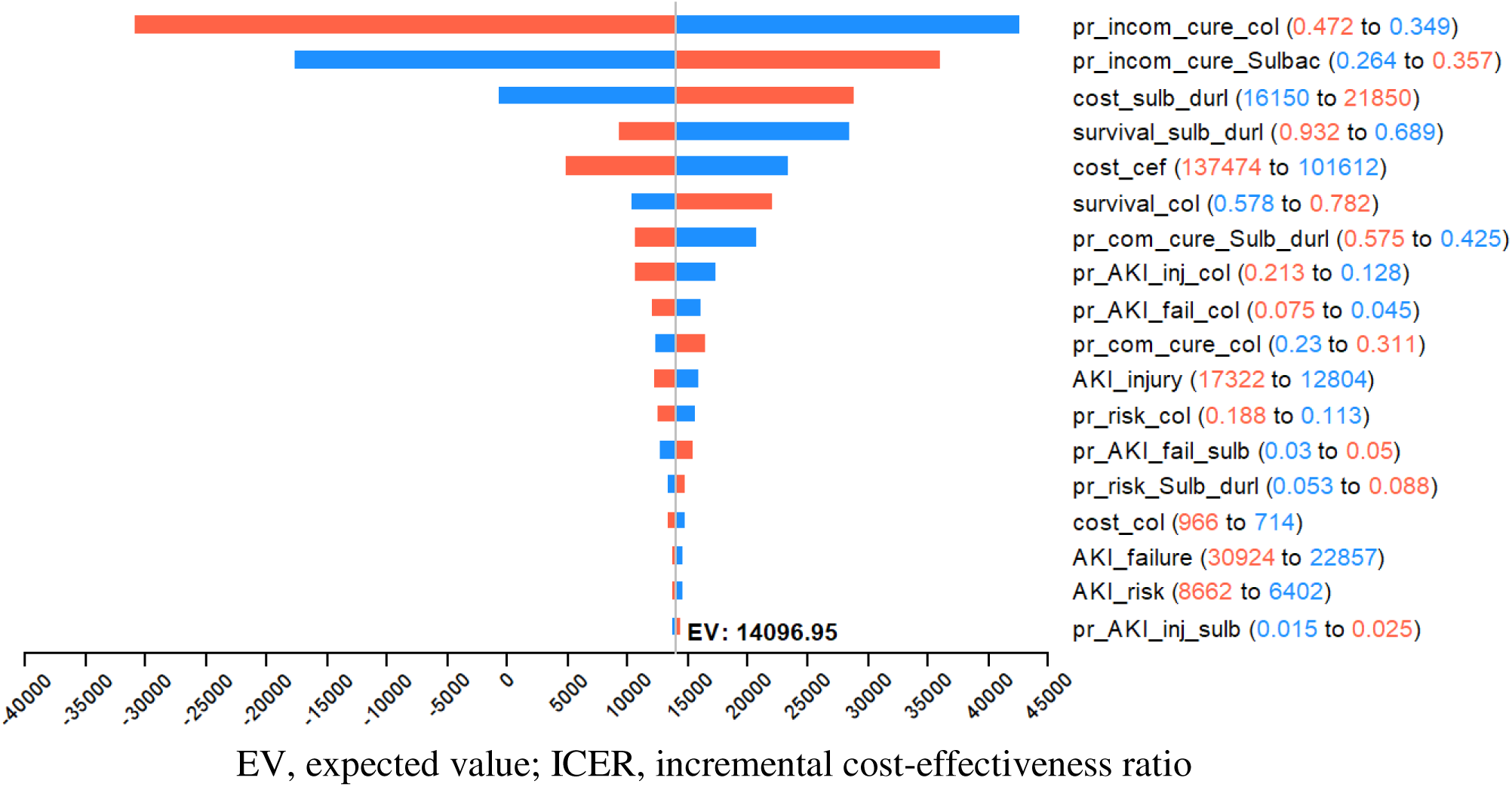
Tornado Diagram ICER (per additional life saved)

### Scenario Analyses

In the first scenario analysis, evaluating survival and clinical cure status at end of therapy (final dosing day), rather than at test of cure, colistin was dominated, with higher costs and no additional benefit. In the second scenario analysis, which utilized clinical cure status at late follow-up, rather than at test of cure, the per-patient cost increased to $75,841 for sulbactam-durlobactam and $62,329 for colistin, resulting in an ICER of $69,758 per additional life saved (Table 3).

**Table 3:**
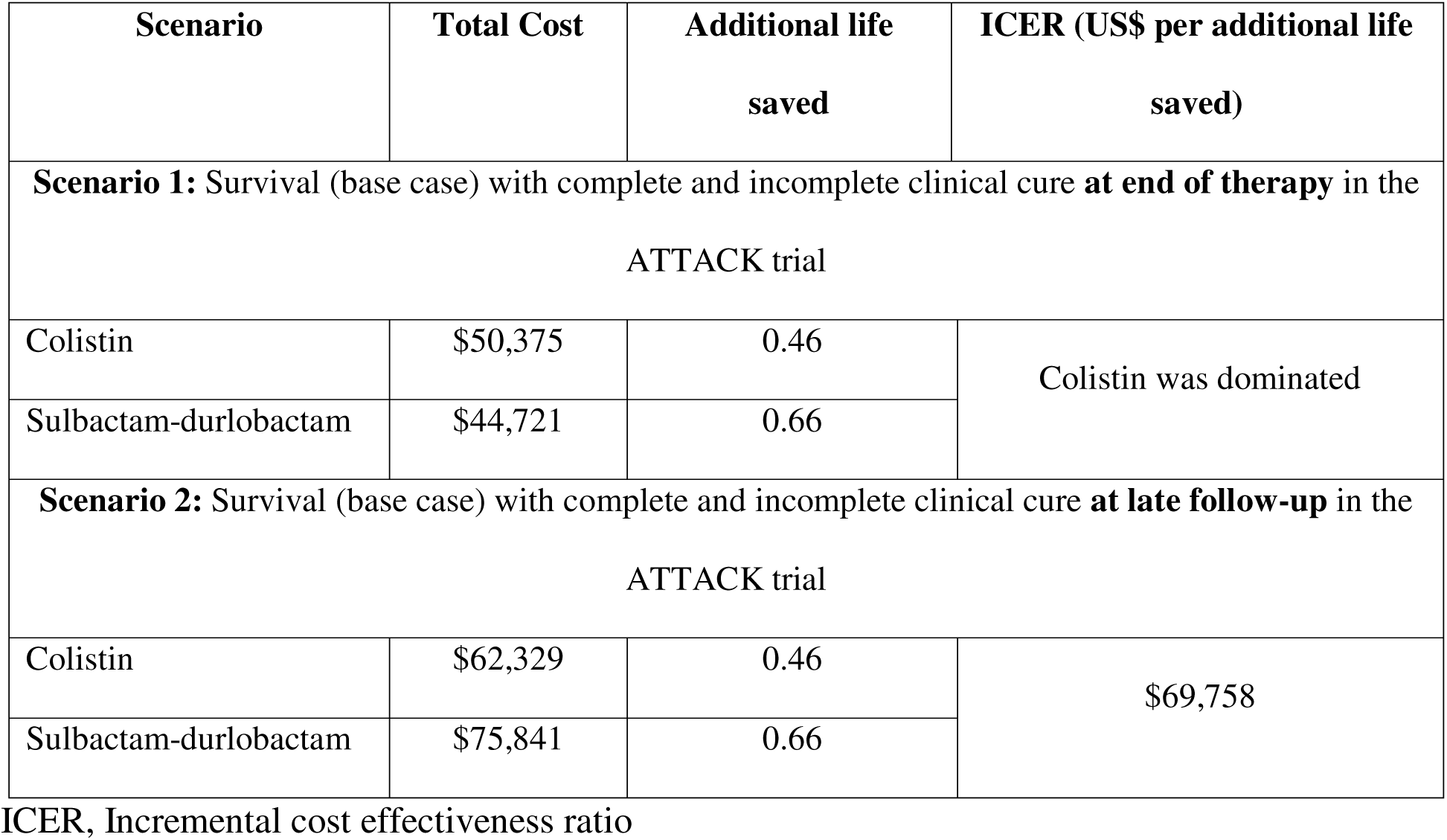
Scenario analyses.

## Discussion

In our study of the cost-effectiveness of sulbactam-durlobactam as compared with colistin for CRAB infections in an inpatient critical care setting in the US, sulbactam-durlobactam incrementally increased both cost (+$2,731) and survival, corresponding to 0.19 additional life saved per patient in the base case, yielding an ICER of $14,097 per additional life saved compared to treatment with colistin.^18^ When extrapolated to a hypothetical cohort of 100 patients admitted with CRAB infections, sulbactam-durlobactam could prevent approximately 19 mortalities compared with colistin, based on the modeled outcomes and absolute rates of cure observed in the ATTACK trial.

Our findings are consistent with prior economic evaluations demonstrating the cost-effectiveness of newer β-lactam/β-lactamase inhibitor therapies relative to colistin, while extending this evidence to sulbactam–durlobactam using trial-based inputs specific to CRAB infections.^8^ That study used a cohort-level decision model informed by meta-analytic effectiveness estimates, whereas our analysis employed a patient-level decision model using trial-based efficacy data from the ATTACK study and incorporated detailed AKI severity stratification and costs specific to CRAB infections.^8^

A 2013 study assessed the economic value of a hypothetical pathogen-specific antibacterial agent for CRAB and reported a net cost per life saved in the US of $15,265 (2013 US dollars), equivalent to approximately $21,531 in 2025 US dollars,^19^ which is higher compared to the cost per additional life saved of $14,097 reported in our study. Our base case findings suggest that sulbactam-durlobactam provides good value when compared with colistin. However, stratified AKI severity and patients surviving with or without complete cure were not assessed in other models. ^8,19^

Scenario analyses highlighted that the economic value of sulbactam-durlobactam was sensitive to key clinical assumptions. Cost-effectiveness was attenuated when alternative treatment sequencing was assumed and was highly dependent on the survival and clinical benefits observed in the ATTACK trial. In contrast, defining treatment success based on clinical cure rather than test-of-cure outcomes favored sulbactam–durlobactam, particularly when response was assessed at the end of therapy. Collectively, these findings indicate that assumptions regarding survival benefit, treatment sequencing, and timing of outcome assessment meaningfully influence modeled cost-effectiveness, suggesting that variability in real-world cure patterns, follow-up practices, and prescribing decisions may affect whether modeled economic value is realized in clinical practice.

These findings are particularly relevant in the context of evolving treatment recommendations for CRAB infections, where clinicians must balance antimicrobial efficacy against toxicity associated with polymyxins when compared to sulbactam-durlobactam in critically ill patients.^7,20^ As newer agents are increasingly incorporated into treatment algorithms, considerations beyond microbiologic activity, such as safety profiles, limited efficacy due to emerging resistance, and downstream consequences, are becoming central to therapeutic decision-making.^7,21^

The explicit inclusion of acute kidney injury-related costs and impact on survival reflects a clinically meaningful driver of morbidity and associated healthcare resource utilization in CRAB treatment.^8,9,19^ Nephrotoxicity remains a well-recognized complication of colistin therapy, and accounting for its impact on cost of treatment underscores the importance of toxicity considerations in economic evaluations of last-line antibiotics.^11,20,22^

Although this analysis leveraged trial-based efficacy and safety data, real-world outcomes may differ due to patient heterogeneity, local resistance patterns, and variability in treatment sequencing and supportive care practices.^11,23^ Such factors may influence both effectiveness and toxicity in routine clinical settings, highlighting the importance of future real-world evaluations to assess the generalizability of these findings.^24^

Our cost-effectiveness study has several limitations. First, the ATTACK trial was designed as a non-inferiority trial, and the base case modeled the treatment difference between sulbactam-durlobactam and colistin, presuming an efficacy difference. Second, the model did not account for the longer term morbidities of AKI. Third, our study did not assess the overall differences in hospital/ICU costs between groups. In addition, our model did not adjust for differences in the duration of hospital stay associated with disease severity and AKI events. Our study modeled a 45-day time horizon based on the ATTACK trial; however, patients in critical care have varying lengths of stay. Fourth, the cost of medication was estimated based on the U.S. wholesale acquisition cost, which might not reflect the actual cost paid by hospitals. Lastly, the cost associated with treating CRAB infections and comorbidities was assumed to be equal in both treatment arms and excluded from the model, yet it may have differed between treatments.

## Conclusion

We evaluated the cost-effectiveness of sulbactam-durlobactam compared with colistin for the treatment of CRAB infections in US critical care settings. Although sulbactam-durlobactam has higher acquisition costs, these differences were attenuated when accounting for the costs of managing acute kidney injury, which occurs more frequently with colistin. Incorporating efficacy differences observed in the ATTACK trial resulted in an ICER of $14,097 per additional life saved. Overall, sulbactam-durlobactam was cost-effective under commonly applied thresholds, driven by improved survival and reduced nephrotoxicity. These findings support the clinical and economic value of sulbactam-durlobactam and may inform antimicrobial stewardship and formulary decision-making. Future studies are needed to validate these results in real-world settings and across diverse hospital systems.

## Declarations

### Ethics declaration

not applicable

### Patient consent for publication

not applicable

### Clinical trial number

not applicable

### Availability of data and materials

The datasets supporting this article are included within the article and are publicly accessible.

### Author Contributions

Conception and design: AO, SK.

Data acquisition, analysis, or interpretation: AO, ARC, SC, SK.

Drafting of the manuscript: AO, ARC, SK.

Critical revision of the manuscript for important intellectual content: AO, ARC, SC, SK.

### Conference Presentation

Part of this work was presented, in part, as a research abstract at the International Society for Pharmacoeconomics and Outcomes Research (ISPOR) 2024 Annual Conference, held from May 5 to 8, 2024 in Atlanta, Georgia.

